# Can Large Language Models Reduce the Cost of Extracting Data from Electronic Health Records for Research?

**DOI:** 10.64898/2026.01.09.26343792

**Authors:** Stuart Hagler, Mohammad Adibuzzaman, Shannon K. McWeeney, Aaron Cohen

## Abstract

**Objective:** Much medical data is only available in unstructured electronic health records (EHR). These data can be obtained through manual (human) extraction or programmatic natural language processing (NLP) methods. We estimate that NLP only becomes economically competitive with manual extraction when there are ~6500 EHRs records. We have found that there is interest from clinicians and researchers in using NLP on projects with fewer records. We examine whether a large language model (LLM) can be used to reduce the cost of NLP to make it economically competitive for such projects, and study the feasibility of such framework for accuracy.

**Materials and Methods:** We developed an NLP pipeline using an off-the-shelf open LLM to extract breast cancer ER, PR, and HER2 biomarker data. Pipeline development stopped when the prompts performances were competitive with manual extraction. The development time and extraction performance were compared to those for an existing rule-based (RB) NLP pipeline. The code for the extraction portion of the LLM pipeline is available at https://github.com/sehagler/llm_biomarker_extraction.

**Results:** The LLM pipeline produced performance competitive with manual data extraction with a hands-on development time that was ~38% that of the RB pipeline.

**Discussion:** LLMs exhibit lower hands-on development costs compared to standard NLP techniques, but require significant and potentially costly computation resources.

**Conclusion:** LLMs may potentially allow the economically competitive application of NLP to smaller projects if computation costs can be managed.

## Introduction

Approximately 80% of healthcare data remain in unstructured form after it is created.[1] These unstructured data have been made available for use by researchers and clinicians largely through the employment of human medical scribes.[2] Natural language processing (NLP) techniques have been explored as a means improve access to unstructured healthcare data by lowering costs and increasing accuracy as compared to medical scribes.[3]

We had developed a rule-based (RB) NLP pipeline capable of extracting data from electronic health records (EHRs) with performance exceeding that typical of manual data extraction (by medical scribes). [4] An RB extraction can be thought as being like a set of regular expressions operating at the word level but which are able to generalize over grammatical or syntactical categories (part-of-speech, whether an item is named entity, etc.). We put the RB pipeline into production at a large research hospital as a fee-based institutional NLP service for researchers and clinicians. We found that the RB pipeline required projects involving at least ~6500 EHRs to be cost effective against manual data extraction. However, we also found that interest in the use of the NLP service came from researchers and clinicians working on smaller projects (e.g. cohort selection, tumor boards, or smaller research studies) for whom the RB pipeline was relatively expensive.

We illustrate the situation in Fig. 1. For a project requiring data be extracted from documents, the total cost of manual data extraction grows linearly with the total number of documents. However, the cost of NLP is approximately constant. There is a large initial cost to prepare a set of training and testing documents and develop NLP tools from them, but once the tools are developed, the cost per document in production is very small. In Fig. 1 the cost of developing NLP tools is *C*\*, and the cost of manual data extraction equals the cost of NLP when there are *N*\* documents. So for projects with more than *N*\* documents, NLP is less expensive otherwise manual data extraction is less expensive. In our situation we estimated *N*\* ~ 6500. As Fig. 1 illustrates, in order to bring down *N*\* to make NLP economical for smaller projects, we must lower *C*\*, the total cost of developing the NLP tools.

**Figure 1.**
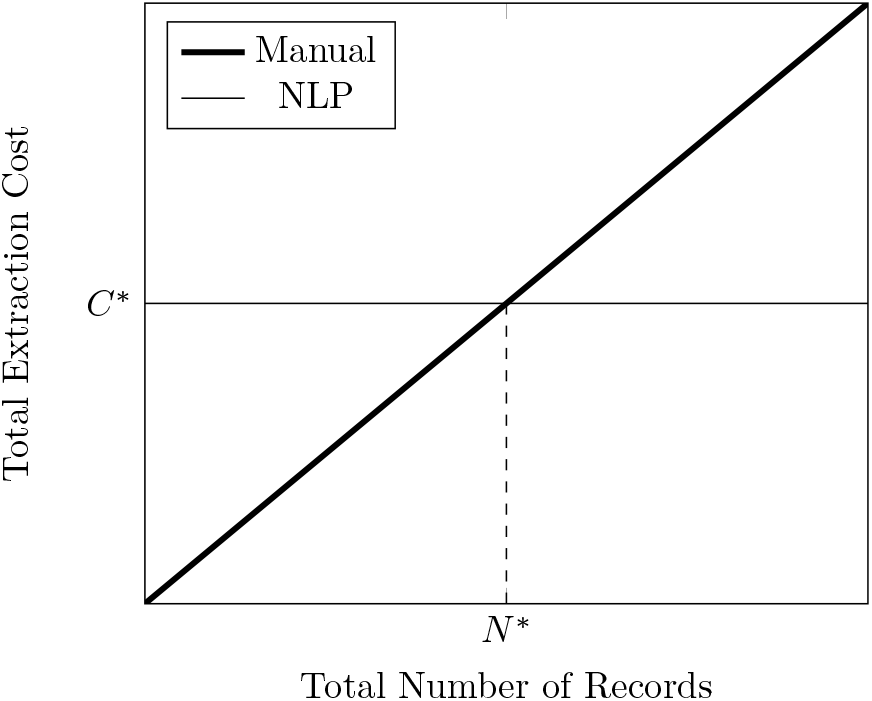
Total extraction cost as a function of total number of records processed for manual extraction and NLP. The total cost of manual extraction rises approximately linearly with the number of records, while NLP has a large initial development cost *C*\* but the additional costs per record processed are small. The number of records at which the two methods have equal cost is *N*\*. NLP is less expensive for number of records greater than *N*\*, while manual review is less expensive when the number of records is less than *N*\*.

Recent advances in large language models (LLMs). [5, 6] have shown the potential to obtain good results versus manual data extraction with much less development effort than was required by earlier NLP techniques. LLMs have been applied to a number of NLP problems in healthcare including question answering, [7, 8, 9] text summarization, [10] and information extraction. [11, 12, 13, 14, 15, 16] This work has been performed using various LLMs including BARD/Gemini,[9] ChatGPT (GPT-3.5 Turbo),[9, 10, 12, 14, 16] Claude 2,[13] GPT-3,[11, 15] GPT-3.5,[8, 9, 10] GPT-4,[8, 9] GPT-JT,[16] Llama-2,[15] and PaLM.[7] LLMs are general language models that must be customized through some additional user input before being put to work on a specific problem. One method of customization is fine-tuning [15] where the model, which has been trained to be a general language model is given further training on a custom data set to teach it how to perform a specific task. Another method is zero-shot prompting [8, 9, 10, 12, 13, 14] where the input presented to the model includes instructions telling the model what to do. Finally a third method is few-shot prompting [7, 11, 14, 16] where the the input presented to the model includes examples of what output the model should produce for a given input. We opted for the few-shot prompting approach as this not only yielded good results, but also because future work could be carried out by trained prompt engineers without expertise in machine learning techniques.

In this project, we begain with a production RB pipeline which used a licensed RB engine to perform RB data extractions. We then replaced the RB engine with an off-the-shelf open-source LLM run in-house on a high-performance computing (HPC) cluster. The aim of the project was *not* to develop NLP data extractions that were competitive with state-of-the-art NLP models. Instead the aim was to see if we could use an LLM to produce results that were (1) competitive with manual data extraction, and (2) having a hands-on development cost (i.e. the cost of the time when the developer is actively working on the project rather than letting the software run unsupervised on some system for a length of time) that was significantly less than that for the existing RB pipeline, while (3) understanding what sorts of technical issues may come up in constructing such a pipeline. The development of software and prompts continued only until the performance of the data extractions was on par with manual data extraction at the end of which the total hands-on development time was recorded.

The actual time taken for each run of the pipeline on the training data was fairly long (upwards of 24 hrs depending on resource availability on the HPC cluster). Each such run has a definite cost for resource utilization that contributes to the overall project cost. Since these resource costs may vary from institution to institution and may be expected to come down in the future, we do not address this cost component in the analysis.

## Materials and Methods

### Study overview

We used an off-the-shelf open-source LLM to construct an NLP pipeline for breast cancer patient EHRs to extract values for 12 variables related to the estrogen receptor (ER), progesterone receptor (PR), and human epidermal growth factor receptor 2 (HER2) biomarkers. The 12 extracted variables consisted of: (1) 4 variables for ER (percentage, score, staining, and strength), (2) 5 variables for PR (percentage, score, staining, strength, and variability), and (3) 3 variables for HER2 (percentage, score, staining). The value of the percentage variables are a numerical percentage, that for the scores is a value from the set {0. 0+, …, 3, 3+}, that for the staining comes largely from the set {equivocal, negative, positive} but also including a number of synonyms, that for the strength largely from the set {moderate, moderate-strong, moderate-weak, strong, weak} also with synonyms occurring, and that for the variability is Boolean taking a value of True when the term {variable} appears appropriately in the text, but False otherwise. We chose these variables a labeled data set was available that we had assembled for use in constructing RB extractions for the existing production RB pipeline allowing us to compare the final performance of the LLM pipeline with that of the RB pipeline. Unfortunately, since the RB pipeline was in production when the RB extractions were developed, records of the time spent developing the extractions were not kept. However, such data were kept for RB extractions in an earlier, pilot, project extracting variables for acute myeloid leukemia (AML). We therefore compare the estimated development time for the LLM pipeline to those recorded for the RB pipeline for AML to arrive at an estimate of how much more quickly extractions may be developed using the LLM.

### RB pipeline

The RB pipeline consisted of 3 components run in sequence: a custom preprocessing pipeline, a licensed NLP engine (Linguamatics I2E), and a custom postprocessing pipeline. The whole sequence was run in-house on a dedicated server running 16 CPUs at 3000 MHz. The preprocessing pipeline took the raw text of an EHR and generate a cleaned and normalized text for ingestion into the NLP engine. The intent of the normalization was to simplify the set of rules used by the NLP engine. The postprocessing pipeline normalized the extractions for an EHR, and performed a final cleaning. When the postprocessor found multiple extractions for the same variable and the rules did not determine a final value, it would flag the variable indicating that a human perform a manual review (MR) of the original EHR to obtain the correct value. The existence of this MR flag in the RB pipeline process is important for our immediate purpose since the data used to train the LLM pipeline included this MR flag as the correct value in some cases.

### LLM pipeline

We constructed the LLM pipeline from RB pipeline by replacing the NLP engine with a module containing an off-the-shelf, open source LLM. The LLM module ingested the normalized documents generated by the preprocessing pipeline and output the extracted data in the same format as the NLP engine. While the preprocessing and postprocessing pipelines continued to be run on the in-house server, the LLM module was run on an in-house HPC cluster containing nodes with Nvidia V100 GPUs.

The LLM module used the Mistral 7B Instruct model [17] which is an off-the-shelf, open source fine-tuning of the Mistral 7B base model,[18] a 7billion-parameter language model. The Mistral 7B Instruct model has been fine-tuned to recognize and follow user-specified instructions. We term this model ‘off-the-shelf’ in the sense that we did not perform any further fine-tuning on it. We prompted the LLM using Language Model Query Language (LMQL), a Python-based programming language for LLM programming with declarative elements. [19, 20, 21, 22] We chose LMQL as it allowed us some user-specified control of the output of the LLM.

The LLM module ingests normalized documents from the preprocessing pipeline. For each document, it searches for all instances of a key (ER, PR, or HER2 depending on which biomarker is being extracted). For each instance of the key, it identifies a neighborhood around the key and checks if a possible value for the variable (percentage, score, staining, strength, or variability depending on which variable is being extracted) is present in the neighborhood. If a possible value is present, a copy of the neighborhood is placed in a prompt and input into to the LLM which returns some value(s). The module then takes the returned value(s) and determines whether they are present in the original neighborhood. Only values that appear in the neighborhood are retained. We originally used neighborhoods to keep LLM processing times down to reasonable lengths, however, we have found the use of neighborhoods also facilitates the extracting multiple values for the same variable when they are present in the document. Finally, the module takes any retained values and checks whether there is surrounding information in the neighborhood which augments the value. For example the value returned by the LLM may be ‘positive’ but a search of the neighborhood determines that ‘positive’ preceded by ‘diffusely’, so ‘diffusely positive’ is the final value.

### Prompts

The neighborhoods are submitted to the LLM using a prompt. We give an example of a prompt function in Fig. 2. This is a Python function named *her2_staining_prompt* which uses LMQL to interact with the LLM. The prompt instructs the LLM to extract staining values for the HER2 biomarker from the neighborhood. The function receives two input values —(1) *prompt_params* which is a dictionary containing a number of static parameters, and (2) *neighborhood* which is the neighborhood. The main body of the prompt consists of 5 components. The first component uses the fact that the Mistral 7B Instruct model has been fine-tuned to recognize text marked up by the tags *[[INST]] and [[/INST]]* as instructions. The second component contains a series of few shot examples indicated by the tag *EXAMPLE:* and the desired outputs for each example indicated by the tag *ANSWER:*. The third component gives the neighborhood indicated by the tag *TEXT:*. The fourth component given by the tag *REASONING:* provides a place for the LLM to put a string giving some kind of “reasoning” for the extraction. This “reasoning” does not reflect how the LLM is actually going about making the extraction, rather it is a string generated by the LLM that reads like reasoning for carrying out the prompt instruction. The generation of this “reasoning” by the LLM seems to result in better results overall. The fifth component given by the tag *ANSWER:* provides a place for the LLM to put its answer (i.e. the returned value). One feature of LMQL is that it provides the user with several ways of constraining the output. In this case, we require the extracted value to be a member of the set {absent, negative, negativity, positive, positivity}.

**Figure 2.**
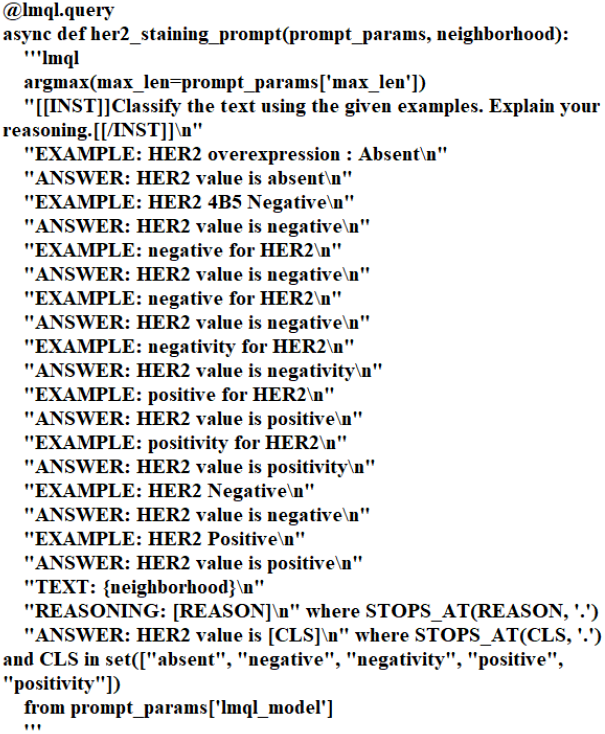
Prompt Function Example.

## Results

### Background

We summarize the relevant observations of a third-party study [23] of the accuracy manual data extraction here for reference. They extracted 27 variables from a data set consisting of 100 unique patient encounters.

The avg. transcription error rate was reported to be 11% (SD, ś21%) per data variable. Thus the avg. accuracy was about 0.89ś0.21. The avg. time for manual data collection per patient encounter was 10ś3.9 min. so the average time for manual data collection per variable was 22ś8.7 sec.

When piloted our RB pipeline using an AML data set. [4] In this pilot study, we developed RB extractions for 5 variables (diagnosis, karyotypes, percentage of blasts in bone marrow, % blasts in peripheral blood, and surface antigens) and recorded their development times in order to estimate the costs for future projects in production. The data set used to develop the queries was divided into training and testing sets. The training set consisted of 134 specimens corresponding to 108 patients, while the testing set consisted of 150 specimens corresponding to 123 patients. No patients appeared in both sets. A complete specimen consisted of two reports —hematopathology and cytogenetics, although some specimens were missing one report. In the training set 27 specimens only had one report present while 107 specimens had both reports, while in the testing set 21 specimens only had one report present while 129 had both reports. There was no flagging for MR in the data in this pilot project.

The trained RB extraction had an avg. accuracy of 0.91ś0.049, an avg. precision of 0.91ś0.072, an avg. recall of 0.91ś0.073, and an avg. F1 of 0.91ś0.071 on the test data set. The avg. time required to develop an RB extraction was ~40 hrs. This includes time for manual extraction of data values in the training and testing sets to produce a gold standard data set. If we assume that manual extraction took about the same time per variable per record given above and that each variable only appeared in one of the two records, then we expect that manual extraction from 284 specimens to have taken ~2 hrs giving an RB extraction development time of ~38 hrs.

We would expect that the manually extracted training and testing data sets would have an accuracy typical of manual review (i.e. 0.89) and that the actual performance of the RB extractions should be adjusted by this amount. However, we have found that the process of RB extraction development involves repeated review of the data sets as extraction errors are investigated resulting in much more accurate training and testing data.

### RB Pipeline on the Breast Cancer Data Set

After the RB pipeline was put into production, we developed extractions to characterize the ER, PR, and HER2 breast cancer biomarkers using 12 variables (ER percentage, ER score, ER staining, ER strength, PR percentage, PR score, PR staining, PR strength, PR variability, HER2 percentage, HER2 score, and HER2 staining). The training data set consisted of 574 patients with 658 (pathology) reports, and a testing set consisting of 660 patients with 771 reports. No patients appeared in both sets. Instances where the extraction should produce a MR flag were indicated in the data set. The combined training and testing sets contained a total of 8645 non-MR cases (i.e. cases where the training or testing value was not an MR flag), with 720ś420 on average per variable, and with a max of 1262 and a min of 54 (for PR variability), and the next lowest number being 1186 (for PR score). They contained a total of 511 MR cases (i.e. cases where the training or testing value was an MR flag), with 43ś11 on average per variable. The accuracies for the RB extractions are compared to the observed avg. manual extraction accuracy of 0.89 in Figs. 3, 4, and 5. The precisions, recalls, and F1-scores for the extractions are reported in Tables 1, 2, and 3 in the columns *RB non-MR* and *RB MR*. The column *RB non-MR* indicates the performance on non-MR cases while *RB MR* indicates performance on MR cases. We estimate the avg. performance of the extractions using the non-MR columns. This gives an avg. accuracy of 0.95ś0.021, an avg. precision of 0.93ś0.038, an avg. recall of 0.93ś0.057, and an avg. F1 of 0.93ś0.044. The RB NLP Engine took 26 min. to perform the data extractions.

**Table 1:** Performance of the RB and LLM pipelines on ER variables. Non-MR indicates performance on non-MR data and MR indicates performance on MR data as described for RB extraction for the breast cancer data set.

| **ER Variable** | **RB**<br>**non-**<br>**MR** | **RB**<br>**MR** | **LLM**<br>**non-**<br>**MR** | **LLM**<br>**MR** |
| --- | --- | --- | --- | --- |
| Percentage - Precision | 0.97 | 0.73 | 0.95 | 0.73 |
| Percentage - Recall | 0.98 | 0.73 | 0.93 | 0.73 |
| Percentage - F1 | 0.97 | 0.73 | 0.94 | 0.73 |
| Score - Precision | 0.90 | 1.0 | 0.89 | 0.0 |
| Score - Recall | 0.88 | 0.5 | 0.88 | 0.0 |
| Score - F1 | 0.89 | 0.67 | 0.88 | NA |
| Staining - Precision | 0.95 | 0.50 | 0.92 | 0.86 |
| Staining - Recall | 0.95 | 0.71 | 0.91 | 0.86 |
| Staining - F1 | 0.95 | 0.71 | 0.92 | 0.86 |
| Strength - Precision | 0.97 | 0.83 | 0.93 | 0.86 |
| Strength - Recall | 0.95 | 0.71 | 0.97 | 0.86 |
| Strength - F1 | 0.96 | 0.77 | 0.95 | 0.86 |

**Table 2:** Performance of the RB and LLM pipelines on PR variables. Non-MR indicates performance on non-MR data and MR indicates performance on MR data as described for RB extraction for the breast cancer data set

| **PR Variable** | **RB**<br>**non-**<br>**MR** | **RB**<br>**MR** | **LLM**<br>**non-**<br>**MR** | **LLM**<br>**MR** |
| --- | --- | --- | --- | --- |
| Percentage - Precision | 0.97 | 0.72 | 0.90 | 0.79 |
| Percentage - Recall | 0.97 | 0.72 | 0.87 | 0.76 |
| Percentage - F1 | 0.97 | 0.72 | 0.88 | 0.77 |
| Score - Precision | 0.91 | 0.60 | 0.85 | 0.75 |
| Score - Recall | 0.96 | 0.23 | 0.95 | 0.23 |
| Score - F1 | 0.94 | 0.33 | 0.90 | 0.35 |
| Staining - Precision | 0.95 | 0.50 | 0.93 | 0.45 |
| Staining - Recall | 0.96 | 0.50 | 0.93 | 0.45 |
| Staining - F1 | 0.96 | 0.50 | 0.93 | 0.45 |
| Strength - Precision | 0.95 | 0.80 | 0.89 | 0.64 |
| Strength - Recall | 0.91 | 0.71 | 0.87 | 0.53 |
| Strength - F1 | 0.93 | 0.75 | 0.88 | 0.58 |
| Variability - Precision | 0.67 | NA | 0.86 | NA |
| Variability - Recall | 0.19 | NA | 0.97 | 0.0 |
| Variability - F1 | 0.30 | NA | 0.91 | NA |

**Table 3:** Performance of the RB and LLM pipelines on HER2 variables. Non-MR indicates performance on non-MR data and MR indicates performance on MR data as described for RB extraction for the breast cancer data set

| **HER2 Variable** | **RB**<br>**non-**<br>**MR** | **RB**<br>**MR** | **LLM**<br>**non-**<br>**MR** | **LLM**<br>**MR** |
| --- | --- | --- | --- | --- |
| Percentage - Precision | 0.85 | 0.71 | 0.92 | 0.70 |
| Percentage - Recall | 0.78 | 0.46 | 0.94 | 0.54 |
| Percentage - F1 | 0.82 | 0.56 | 0.93 | 0.61 |
| Score - Precision | 0.93 | 0.80 | 0.84 | 0.80 |
| Score - Recall | 0.94 | 0.73 | 0.84 | 0.73 |
| Score - F1 | 0.94 | 0.76 | 0.84 | 0.76 |
| Staining - Precision | 0.90 | 0.11 | 0.88 | 0.10 |
| Staining - Recall | 0.92 | 0.10 | 0.90 | 0.10 |
| Staining - F1 | 0.91 | 0.11 | 0.89 | 0.10 |

**Figure 3.**
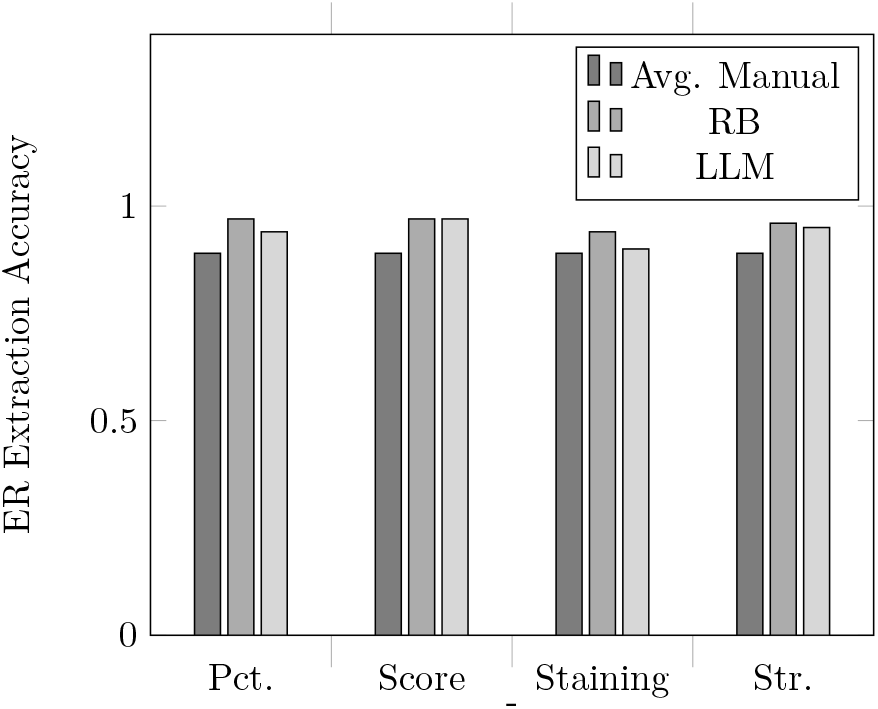
Accuracy of the RB and LLM pipelines on ER percentage, score, staining, and strength variables. The avg. manual extraction value of 0.89 is provided for reference.

**Figure 4.**
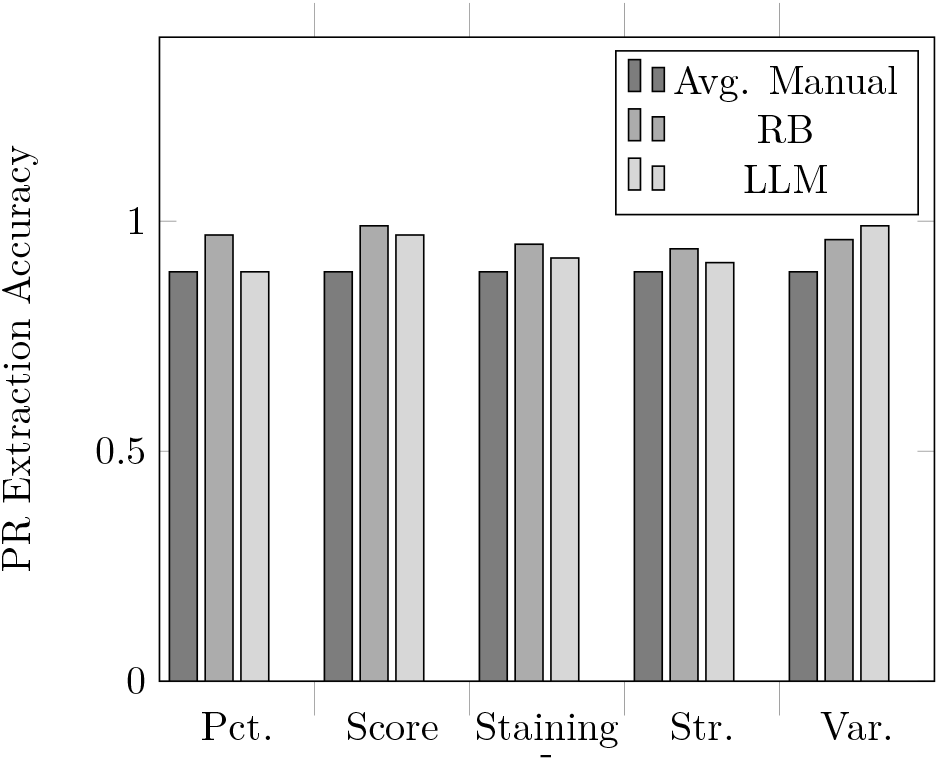
Accuracy of the RB and LLM pipelines on PR percentage, score, staining, strength, and variability variables. The avg. manual extraction value of 0.89 is provided for reference.

**Figure 5.**
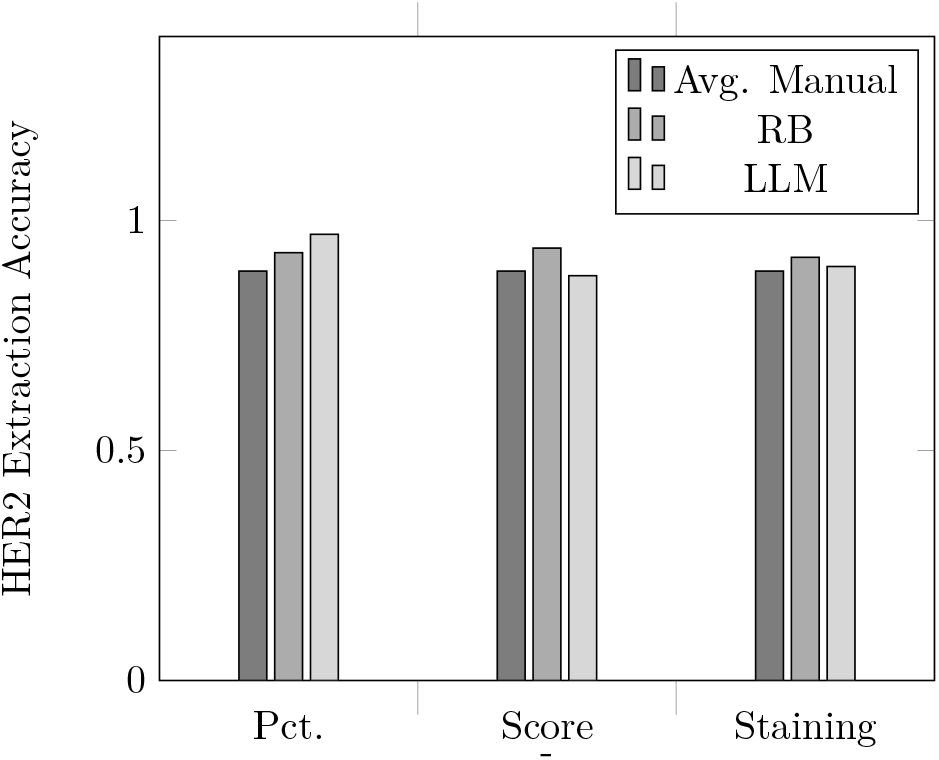
Accuracy of the RB and LLM pipelines on HER2 percentage, score, and staining variables. The avg. manual extraction value of 0.89 is provided for reference.

### LLM Pipeline on the Breast Cancer Data Set

While the RB pipeline was in production we found that we received interest in the RB pipeline from clinicians and researchers involved in smaller projects. We piloted an LLM pipeline using the Breast Cancer data set developed for the RB pipeline to see if that pipeline could operate more cheaply and accommodate smaller projects. The accuracies for the LLM extractions are compared to the avg. manual extraction accuracy of 0.89 [23], and RB extractions in Figs. 3, 4, and 5. The precisions, recalls, and F1-scores for the extractions are reported in Tables 1, 2, and 3 in the columns *LLM non-MR* and *LLM MR*. We again estimate the avg. performance of the extractions using the non-MR columns. This gives an avg. accuracy of 0.93ś0.037, an avg. precision of 0.90ś0.035, an avg. recall of 0.91ś0.042, and an avg. F1 of 0.90ś0.032. The total number of few-shot examples was 186 giving an avg. of 15.5 prompts per variable. The total hands-on time spent developing the extractions was ~160 hrs or ~13 hrs of development time for each variable. We estimate that approximately half of the development time was spent developing the prompts with the other half spent developing the Python module wrapping the LLM. The development of the extractions took place over a period of 4 months. We divided the extractions among three scripts —one running the ER extractions, a second running the PR extractions and a third running the HER2 extractions. The scripts were run on a node of an HPC cluster allocated with 4 V100 GPUs and 20 CPUs. Each script ran in ~24 hrs, so the extractions could generally be run in ~24-72 hrs depending on resource availability on the HPC cluster.

## Discussion

Our aim has been to develop an LLM pipeline capable of performing extractions at a significantly lower cost than an existing RB pipeline but having a performance that is competitive with manual data extraction. We estimated that the RB pipeline required ~40 hrs to develop extractions that performed at least as well as manual data extraction. For the LLM pipeline, the extraction the hands-on development of one variable required ~13 hrs of development if labeled testing and training data were available, and ~15 hrs if they were not. In the latter case, we estimate that the hands-on development time, and thus cost, of the LLM pipeline was ~38% that of the RB pipeline. As we indicated in the introduction, we are not addressing the cost of the computation resources as these may vary from institution to institution and may be expected to come down in the future, but this does contribute to the overall cost of LLM pipeline development.

The ~13 hrs of development time for the module for each LLM extraction of one variable consisted of about half the time being spend on developing the prompt and half the time developing the Python code in the module. This project was a pilot rather than a production project and involved some amount of trial and error to obtain extractions with the required performance. In a mature, production LLM pipeline, we expect that the developer would be able to reduce the Python wrapper code into a package of reusable functions. In this case the development time would be expected to be lower.

As we have mentioned, there is a cost associated with utilizing the computation resources needed to run the LLM pipeline. During prompt development, the LLM pipeline was run in two modes. In the first mode, the LLM pipeline would be run on a single record usually using a single prompt to debug a specific issue. In this mode, the LLM pipeline could be run on a laptop or on the dedicated servers for the RB pipeline and did not accrue an additional cost. In the second mode, the LLM pipeline was run over the full set of training data to measure the current performance of one or more prompts. In this mode the LLM pipeline was typically run on an HPC cluster with base usage fee and further fees based on resource usage. In our development process, we found that we were not always able to get the desired GPU resources on the HPC cluster. To maintain a steady development timeline we ran individual biomarker prompt sets (i.e. the ER, or PR, or HER2 prompts run as a group) on either the HPC cluster using only CPUs or on the dedicated servers for the RB pipeline. While the run-time was longer than with GPUs, the time was still reasonable. The prompt developer does have the option of accepting longer times for each run of the LLM pipeline in exchange for lower computation costs.

The prompts extracted values well there was a correct value to be extracted in the neighborhood and that value occurred in a context that was a reasonable generalization from the few-shot examples. Even when multiple values of the same type were present (e.g. multiple percentage values), the LLM generally returned the correct value. The main problem we encountered was when the neighborhood did not contain a correct value to be extracted. These cases divided into two types: (1) where there was no value in the neighborhood corresponding to what the prompt was looking for, the LLM frequently hallucinated a value, and (2) where there was one or more values in the neighborhood corresponding to what the prompt was looking for, the LLM frequently returned one of these other values. Cases of the first type were easily dealt with by the simple expedient of programmatically confirming whether any value extracted by the LLM was actually present in the neighborhood. Cases of the second type were more difficult to handle and we adopted a strategy of judiciously selecting the neighborhoods to avoid this problem as much as possible, and to do some programmatic postprocessing to handle the more frequent errors of this type. We do feel that this strategy results in code outside of the prompt is doing more heavy-lifting than we would like, and that more robust solutions involving the LLM should be researched.

Finally, the prompts exhibited a certain fragility. We found that small changes in the wording of the prompts would change the performance of the LLM even when the changes looked to a native speaker as amounting to the same thing. Generally, small changes to a prompt generated 2 or 3 percentage point changes in the accuracy, precision, recall, or F1 values, although occasionally much larger changes in performance were observed. We dealt with these issues by making some decisions early on and holding to them throughout development. For example, the layout of the prompt, the text of the instruction, and the style of the few-shot examples were the result of early decisions. This fragility very likely means that prompts will exhibit changes in performance when transferred to a different LLM or a newer iteration of the LLM the were written for.

## Conclusion

Standard NLP techniques are too expensive to use on smaller clinical and research projects while at the same time there is interest in the use of such techniques from clinicians and researchers. LLMs exhibit lower hands-on development costs compared to standard NLP techniques. However, LLMs require significant computation resources which add a further cost to the development and deployment of LLM techniques. If these computation costs can be managed and kept sufficiently low then LLMs have the potential to make NLP accessible to smaller projects in an economically competitive way.

## Data Availability

All data produced in the present work are contained in the manuscript.

## Acknowledgments

The authors thank Steven D. Bedrick for advice on experimental design.

